# Detection and prevalence of SARS-CoV-2 co-infections during the Omicron variant circulation, France, December 2021 - February 2022

**DOI:** 10.1101/2022.03.24.22272871

**Authors:** Antonin Bal, Bruno Simon, Gregory Destras, Richard Chalvignac, Quentin Semanas, Antoine Oblette, Gregory Queromes, Remi Fanget, Hadrien Regue, Florence Morfin, Martine Valette, Bruno Lina, Laurence Josset

## Abstract

In Dec 2021-Feb 2022, an intense and unprecedented co-circulation of SARS-CoV-2 variants with high genetic diversity raised the question of possible co-infections between variants and how to detect them. Using 11 mixes of Delta:Omicron isolates at different ratios, we evaluated the performance of 4 different sets of primers used for whole-genome sequencing and we developed an unbiased bioinformatics method which can detect all co-infections irrespective of the SARS-CoV-2 lineages involved. Applied on 21,387 samples collected between weeks 49-2021 and 08-2022 from random genomic surveillance in France, we detected 53 co-infections between different lineages. The prevalence of Delta and Omicron (BA.1) co-infections and Omicron lineages BA.1 and BA.2 co-infections were estimated at 0.18% and 0.26%, respectively. Among 6,242 hospitalized patients, the intensive care unit (ICU) admission rates were 1.64%, 4.81% and 15.38% in Omicron, Delta and Delta/Omicron patients, respectively. No BA.1/BA.2 co-infections were reported among ICU admitted patients. Although SARS-CoV-2 co-infections were rare in this study, their proper detection is crucial to evaluate their clinical impact and the risk of the emergence of potential recombinants.

## Introduction

Since the first SARS-CoV-2 genome was published in January 2020, five variants of concern (VOC), characterized by increased transmissibility and/or immune escape capacity, have circulated worldwide^1^. Omicron, the last VOC to date, was first detected in South Africa^2^ in November 2021 and displaced the previously circulating Delta VOC by February 2022. While the Delta and Omicron variants share a few mutations along their genome, Omicron is characterized by a large number of specific mutations, especially in the S gene^3^. In December 2021, a sub-lineage of the Omicron variant, named BA.2, was detected and co-circulated with the Omicron BA.1 lineage from January 2022. Although derived from a common ancestor (lineage B.1.1.529), BA.1 and BA.2 are highly divergent with 27 specific mutations for the latter ^3^.

In France, the fifth wave of the SARS-CoV-2 epidemic was characterized by a sustained co-circulation of the Delta and Omicron (lineage BA.1) variants from November 2021-January 2022. Lineage BA.2 was first detected in France in late December 2021 and its proportion has since increased linearly ^4^. Thus, the unprecedented sustained co-circulation of genetically divergent lineages observed from November 2021 to February 2022 may have been suitable for co-infections with a risk of subsequent recombination events. Few cases of Delta and Omicron variant co-infections have been reported recently but a systematic assessment of the prevalence of SARS-CoV-2 co-infections, including BA.1/BA.2 co-infections, has not been explored on a large data set ^5–7^. As Omicron and Delta have been associated with distinct COVID-19 severity, the clinical spectrum of these co-infections also needs to be determined ^8,9^.

Detection of SARS-CoV-2 co-infection using whole-genome sequencing (WGS) is not trivial and can be hampered by several problems: i) uneven performances of primer sets used to amplify SARS-CoV-2 genome with possible amplification bias of some genomic regions of specific variants ^10,11^; ii) possible sample contamination during the sequencing process which requires independent validation on duplicate extracts ^12,13^; and iii) lack of unbiased and validated bioinformatics methods able to systematically detect co-infections. Previously published reports on SARS-CoV-2 Delta/Omicron co-infections were based on manually curated lists of divergent mutations and visual examination of their relative frequencies along the genome ^5–7^. De-novo assembly methods have also been described to assemble different viral genomes present in one sample ^14^ but are computationally intensive.

Herein, we used different mixed ratios of Delta and Omicron cell culture isolates in order to assess the performances of four different primer sets for the detection of Delta/Omicron co-infections. A co-infection score was determined to warn about probable co-infection. The prevalence of Delta/Omicron and BA.1/BA.2 co-infections were then estimated on a large data set of sequences obtained from random surveillance of out-patients and systematic sequencing of hospitalized patients.

## Methods

### Mix of Delta:Omicron cell culture isolates

The Delta and Omicron variants were isolated in cell culture from nasopharyngeal swabs (NPS). Following interim biosafety guidelines established by WHO, NPS were inoculated on confluent Vero E6 TMPRSS2 cells with DMEM supplemented with 2% penicillin– streptomycin, 1% L-glutamine, 2% G418 and 2% inactivated fetal bovine serum. Plates were incubated at 37 °C with 5% CO_2_ for 48 h. The cytopathic effects were monitored daily; samples were harvested when positive. Viral isolates were quantified using RT-PCR ^15^ and sequenced to confirm the lineage and the absence of low frequency diversity. The Delta and Omicron isolates were then diluted to reach similar viral loads (Ct = 19) and mixed using different Delta:Omicron ratios: 0:100, 10:90, 20:80, 30:70, 40:60, 50:50, 60:40, 70:30, 80:20, 90:10 and 100:0. After nucleic acid extraction was performed in duplicate, all RNA extracts were diluted ten-fold and stored in several aliquots under the same conditions (frozen at - 80°C). Thus, all extracts were subjected to one freeze-thaw cycle for all sequencing methods.

### SARS-CoV-2 sequencing

RNA from culture supernatants was extracted in duplicate using the automated MGISP-960 workstation using MGI Easy Magnetic Beads Virus DNA/RNA Extraction Kit (MGI Tech, Marupe, Latvia) for the first extract and EMAG platform (bioMérieux, Lyon, France) for the second extract. A total of four sets of SARS-CoV-2 primers (Artic V4 and V4.1, Midnight V1 and Midnight V2) were tested on duplicate extracts of culture supernatants. For Artic V4 and V4.1 primers, cDNA synthesis and amplification were performed using the COVIDSeq-Test™ (Illumina, San Diego, USA). For Midnight V1 and V2 primers, cDNA synthesis and amplification were performed using LunaScript® and Q5® Hot Start (New England Biolabs, Ipswich, USA), respectively. Libraries were prepared with the COVIDSeq-Test (Illumina, San Diego, USA), and samples were sequenced with 100 bp paired-end reads using the NovaSeq 6000 Sequencing system SP flow cell.

Routine SARS-CoV-2 sequencing protocol in our laboratory is based on COVIDSeq-Test™ (Illumina, San Diego, USA) using Artic V4 or V4.1 primers as they became available.

### Sample selection

The samples sequenced at the National Reference Center (NRC) of Respiratory Viruses of Hospices Civils de Lyon (HCL) selected for this study were i) samples from systematic sequencing of hospitalized patients in the Lyon area (university hospital of Lyon, HCL) and from HCL health care workers; ii) samples from random sequencing performed during the weekly Flash survey conducted by the EMERGEN consortium (French consortium for the genomic surveillance of emerging pathogens). The Flash surveys are nationwide surveys where all private and public diagnostic laboratories in France are asked to provide to the NRC and other sequencing centers a fraction of positive samples from one day per week ranging from 25% to 100% according to the number of positive cases detected at the national level ^4,16^. The prevalence of SARS-CoV-2 co-infections was estimated on samples collected both in the HCL and in Flash samples sequenced by the NRC of HCL. To assess the clinical presentations of co-infected patients, three groups were selected: out-patients of Flash surveys, hospitalized patients of Flash surveys and HCL, and healthcare workers of HCL, excluding follow-up samples.

### Bioinformatics

Reads were processed using the in-house bioinformatic pipeline seqmet (available at https://github.com/genepii/seqmet, software versions provided in **Table S1**). Paired reads were trimmed with cutadapt to remove sequencing adapters and low-quality ends, only keeping reads longer than 30 bp. Alignment to the SARS-CoV-2 reference genome MN908947 was performed by Minimap2. Mapped reads were processed to remove duplicates tagged by picard, then realigned by abra2 to improve indel detection sensitivity and finally clipped with samtools ampliconclip to remove read ends containing primer sequences. Variants present at frequencies of 5% or above were called using freebayes, then decomposed and normalized with vt and filtered with bcftools to eliminate false positives. To detect co-infection, obtained vcf files were compared to a lineage variant database, both developed internally to this end. The database consists of vcf files listing variants found in 50% or more of 100 randomly selected sequences for a given pangolin lineage in the full GISAID dataset available (extracted on 02 February 2022). The database is available at https://github.com/genepii/seqmet. The co-infection detection script searches for each lineage any major or minor variant matching expected variants of the putative main lineage and then searches for any minor variant matching any other lineage excluding variants in common with the main lineage. This approach provides putative main and secondary lineages contained in a sequenced sample read, along with a ratio of observed and expected variants in each case. Variants were expected when they occurred on a position covered with at least 100 reads. Secondary lineages, considered indicative of a putative co-infection, required at least 6 lineage-specific variants to be called.

For Delta:Omicron mixes, vcf files were also analyzed using the curated list of clade-defining mutations of Delta and Omicron (lineage BA.1) as previously published ^5,6^. This list is based on https://covariants.org/variants as of 11/02/2022, excluding 21846 C>T (S: T95I) which is present in 40% of Delta variants.

### Statistical analysis

Continuous variables are presented as means ± standard deviations (SD) or median with interquartile range (IQR) and compared using non-parametric Kruskal–Wallis or Mann– Whitney tests. Proportions were compared using the chi-squared or Fisher’s exact test, as appropriate. A p value of <0.05 was regarded as statistically significant. Statistical analyses were conducted using R software, version 4.0.5 (R Foundation for Statistical Computing).

### Ethics

Samples used in this study were collected as part of an approved ongoing surveillance conducted by the NRC of HCL. The investigations were carried out in accordance with the General Data Protection Regulation (Regulation (EU) 2016/679 and Directive 95/46/EC) and the French data protection law (Law 78–17 on 06 January 1978 and Décret 2019–536 on 29 May 2019). Samples were collected for regular clinical management, with no additional samples for the purpose of this study. Patients were informed of the research and their non-objection approval was confirmed. This study was approved by the ethics committee of the Hospices Civils de Lyon (HCL), Lyon, France and registered on the HCL database of RIPHN studies (AGORA N°41).

## D***ata availability***

The GISAID accession numbers of the Delta and Omicron virus isolates used for experimental mixes are EPI_ISL_11171170 and EPI_ISL_11171169, respectively. Sequencing data of the Delta:Omicron mixes were deposited on the SRA database under accession PRJNA817870, and dehosted sequencing data of NPS with co-infections were deposited under accession PRJNA817806.

## Results

### Evaluation of 4 different primer sets to detect SARS-CoV-2 co-infections using WGS

To simulate co-infections, Delta (B.1.617.2) and Omicron (BA.1) viral isolates were mixed using different Delta:Omicron ratios: 0:100, 10:90, 20:80, 30:70, 40:60, 50:50, 60:40, 70:30, 80:20, 90:10 and 100:0. Resulting mixes had fixed viral loads (median Ct = 22.8; IQR=1.5). Four sets of primers (Artic V4 and V4.1, Midnight V1 and V2) were used in duplicate on extracts to test the impact of PCR amplification prior to sequencing on co-infection characterization. All mixes were sequenced to 1 M paired-end reads leading to SARS-CoV-2 genome covered > 98% with median coverage of 2276X (IQR=315X) (**Table S2)**.

The evaluation of the primer sets was performed using a previously published method based on a curated list of mutations specific to Delta and to Omicron derived from co-variants ^5–7^ (**Table S2 and Fig 1**). More than 90% of the Delta-specific mutations were found in all mixes with the 4 primer sets. In contrast, the detection rate of Omicron-specific mutations ranged from 27% in the 90:10 mix using Midnight primers (V1 and V2) to >78% for mixes with expected frequency of Omicron above 30% (**Fig 1A and Table S1**). Medians of covered allele frequency for the specific mutations were used to estimate viral frequency. Relations between measured and expected frequency were not linear (**Fig 1B**). Over-estimation of Delta was observed for all mixes with all primer sets, and especially in mixes with expected frequency of Delta under 30% and sequenced with Midnight primer sets. Measured frequencies of Delta for the 10:90 mix were between 30-33% with Midnight V1 and V2, and 21-25% with Artic V4 and V4.1.

**Figure 1:**
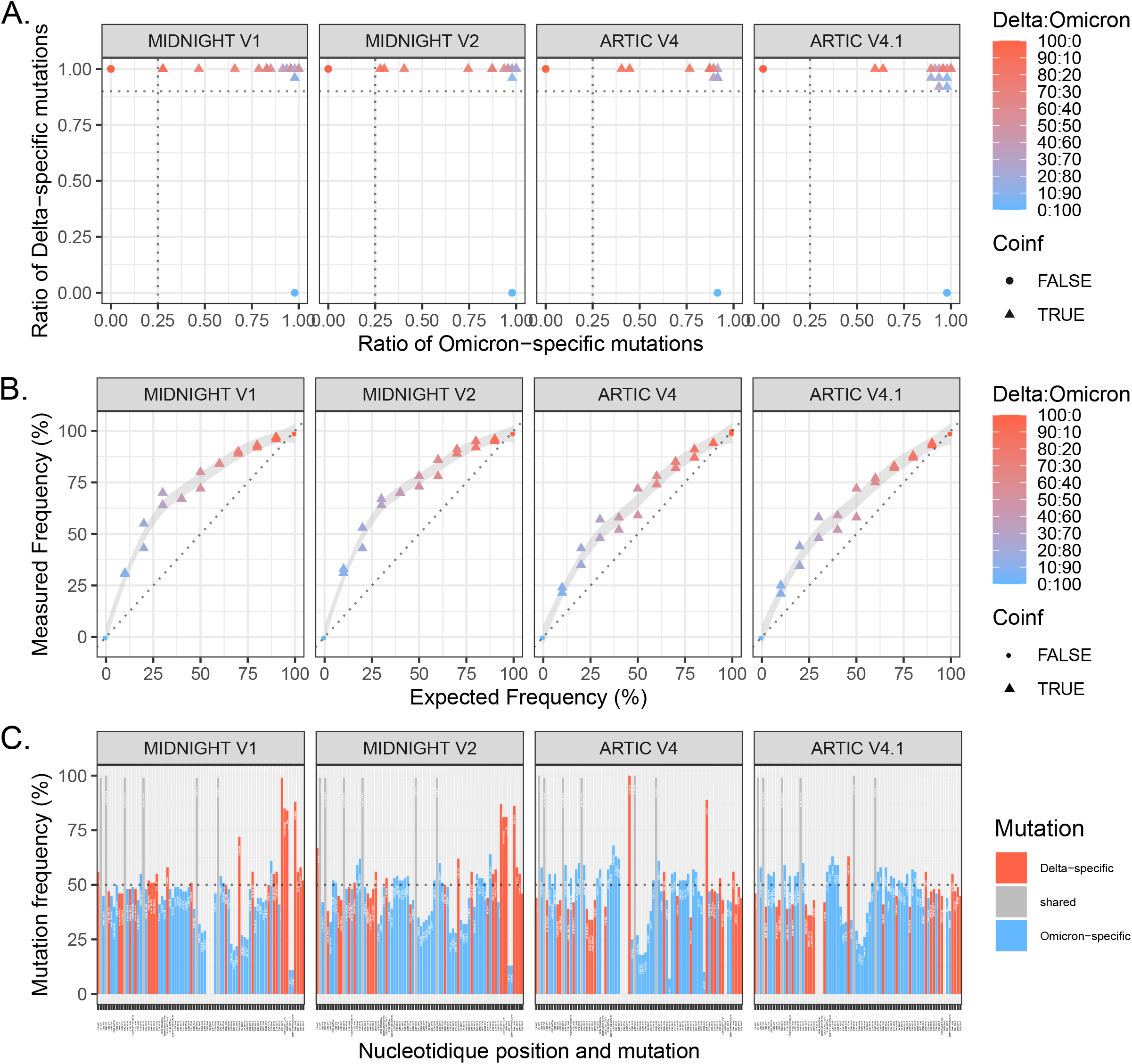
Evaluation of 4 primer sets for WGS of Delta:Omicron mixes: Midnight V1, Midnight V2, ARTIC V4 and ARTIC V4.1. Eleven mixes of Delta:Omicron isolates with different proportions were extracted and sequenced in duplicate. A. Ratio of specific mutations (as minor or major allele) detected from a list of Delta-specific mutations and Omicron-specific mutations based on covariant.org. Horizontal line (Delta-specific mutation ratio) at 0.9 and vertical line (Omicron-specific mutation ratio) at 0.25 discriminate co-infections from pure isolates. B. Frequency of the Delta variant was measured using the median of covered allele frequencies of Delta-specific mutations and compared with expected relative frequency in each mix. Pure isolates (denoted as 100:0 for Delta and 0:100 for Omicron) are depicted with dots, and Delta:Omicron mixes with triangles. Colors represent the expected frequency of each mix using a red (Delta: 100:0) to blue (Omicron: 0:100) gradient. C. Representation of variant allele frequency of Delta- and Omicron-specific mutations for the mix 20:80. Delta-specific mutations are depicted in red, Omicron-specific mutations in blue and shared mutations are in grey. Horizontal lines at 50% show which mutations are called in the consensus sequence based on the majority rule. Chimeric sequences with both Delta and Omicron mutations were detected for the 4 primer sets.

Importantly, consensus sequence calling based on majority rule resulted in artefactual chimeric Delta-Omicron sequences for several mixes and with different patterns depending on the primers used for amplification (**Fig S1**). Chimeric sequences were observed with the four primer sets in all duplicates only for the 20:80 mix (**Fig 1C)**. With Artic primer sets, chimeric sequences were characterized by Omicron sequences bearing the S:L452R and M:I82T mutations, and additional Delta-specific mutations with increasing Delta concentration. With Midnight primers, chimeric sequences were characterized by Omicron sequences with 3’ end of the genome belonging to Delta (starting from nt 27638 or 27874).

Altogether, the Artic V4.1 primers were the least biased for Delta/Omicron co-infection detection and relative frequency estimation, but all primer sets could lead to artefactual chimeric sequences, highlighting the importance of proper co-infection detection.

### Novel bioinformatic algorithm to detect SARS-CoV-2 co-infections using WGS

Independent to this specific set of mutations, an agnostic approach was developed to detect co-infections regardless of the lineage present in the sample (**Fig 2**). This approach is based on the identification of a potential secondary lineage, after excluding variants shared with the main lineage. A secondary lineage is identified only if 6 specific mutations are present. Two ratios are calculated: the main lineage mutation ratio and the secondary lineage mutation ratio quantifying the fraction of present mutations among covered specific mutations. Based on this co-infection detection script, co-infection was successfully identified in all mixes, except for the pure Delta (100:0) and Omicron (0:100) isolates for which only Delta (lineage B.1.617.2) and Omicron (lineage BA.1) were identified as the main lineages, respectively. B.1.617.2 and BA.1 were identified as the main and secondary lineages, respectively, in all mixes with expected frequency of Delta above 40%, independent of the primer sets (**Fig 2 and Table S1**). Main lineage mutation ratios were above 0.9 for all mixes (**Fig 2A)**. Secondary lineage mutation ratios were between 0.216 and 0.941 (**Fig 2B**). The lowest ratios were found for the mix 90:10 with only 0.23, 0.25, 0.39 and 0.55 of BA.1 specific mutations found using Midnight V2, Midnight V1, Artic V4 and Artic V4.1, respectively.

**Figure 2:**
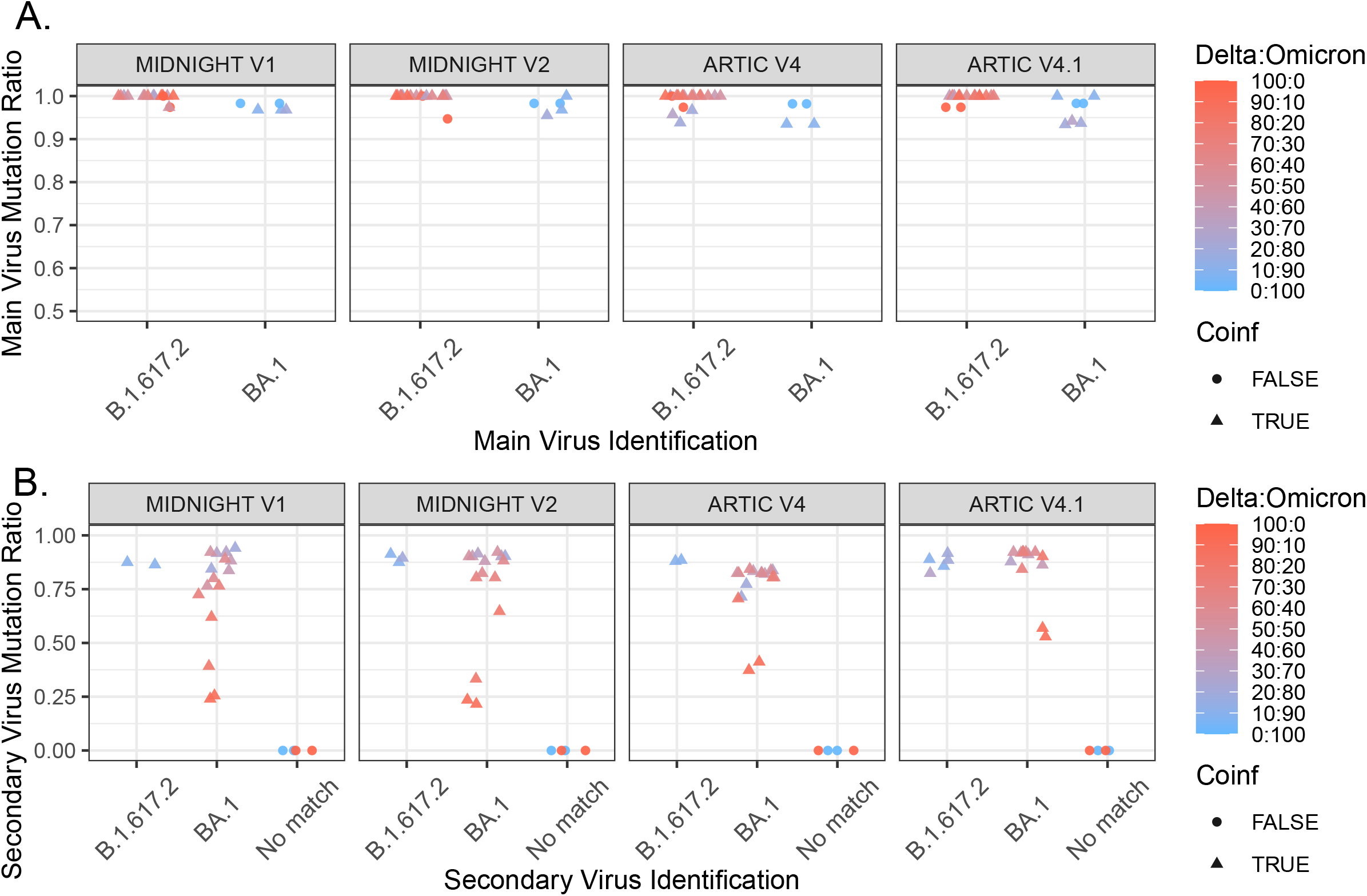
Best main and secondary lineage matches identified for each Delta:Omicron mixes, based on without a priori comparison to a comprehensive lineage mutation database. A. Best main lineage match ratio for each Delta:Omicron mix, considering all mutations expected for the given lineage. B. Best secondary lineage match ratio for each mix, considering mutations expected for the given lineage excluding those in common with the main lineage. Pure isolates (denoted as 100:0 for Delta and 0:100 for Omicron) are depicted with dots, and Delta:Omicron mixes with triangles. Colors represent the expected frequency of each mix using a red (Delta: 100:0) to blue (Omicron: 0:100) gradient.

Altogether, the results of the unbiased co-infection detection scripts were consistent with the curated list approach with a better detection of the secondary lineage with Artic primers.

### Prevalence of SARS-CoV-2 co-infections during the fifth wave in France

Between December 6th 2021 (week 49-2021) and February 27th 2022 (week 08-2022), 23,242 samples were sequenced using Artic V4 or V4.1 primer sets as they became available. In total, WGS (coverage >90%) were obtained for 21,387 samples collected from Flash surveys (n=16,220) and from HCL and peripheral hospitals (n=5,167).

Among the 21,387 samples, 64 samples (0.30%) had a secondary lineage identified with positive ratios (**Fig 3)**. To rule out potential contamination during initial sequencing, all these 64 samples were re-extracted and sequenced in duplicate. In total, 53 samples had a positive secondary lineage mutation ratio in duplicate: 28 samples were identified as a Delta/Omicron (BA.1) co-infection; 1 sample was identified as a Delta/Omicron (BA.2) co-infection; 24 samples were identified as a co-infection between 2 different Omicron lineages (BA.1 and BA.2) (**Fig 4**).

**Figure 3:**
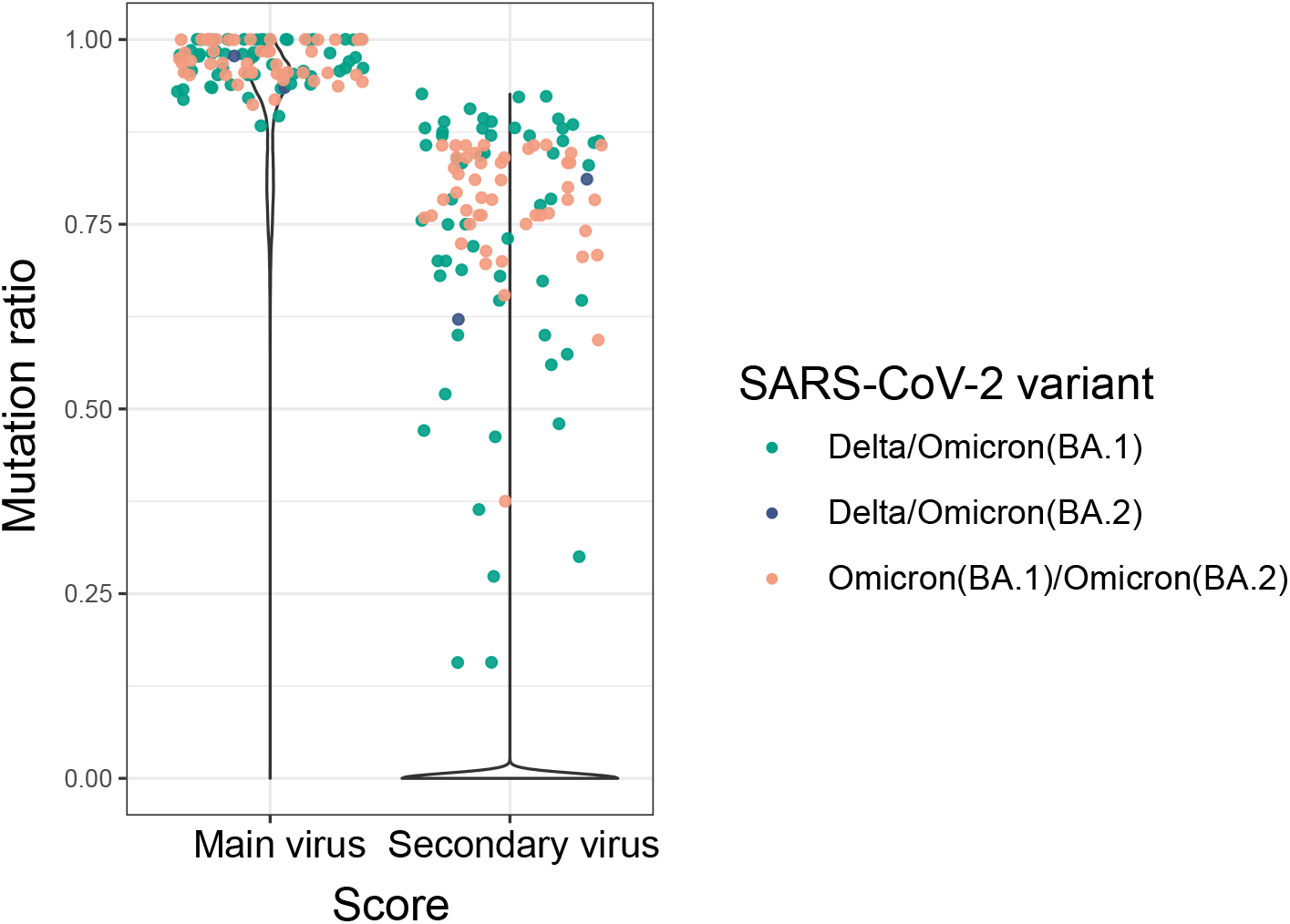
Violin plot of main and secondary virus mutation ratios for the 21,387 samples, including 64 samples sequenced in duplicate. Lineage ratio is considered to be 0 in cases where less than 6 mutations were found or covered with at least 100x for any lineage. Positive secondary virus mutation ratio in independent duplicates with concordant virus identification were considered as potential co-infections and depicted with dots (n=53). Colors indicate lineages : Delta (B.1.617.2 and AY.* lineages) in red, Omicron (BA.1) in blue, Omicron (BA.2) in purple, Omicron (BA.3) in light blue, other lineages (including B.1.640.1) in grey. Co-infections between Delta/BA.1 are in green, Delta/BA.2 in dark blue, and BA.1/BA.2 in salmon.

**Figure 4:**
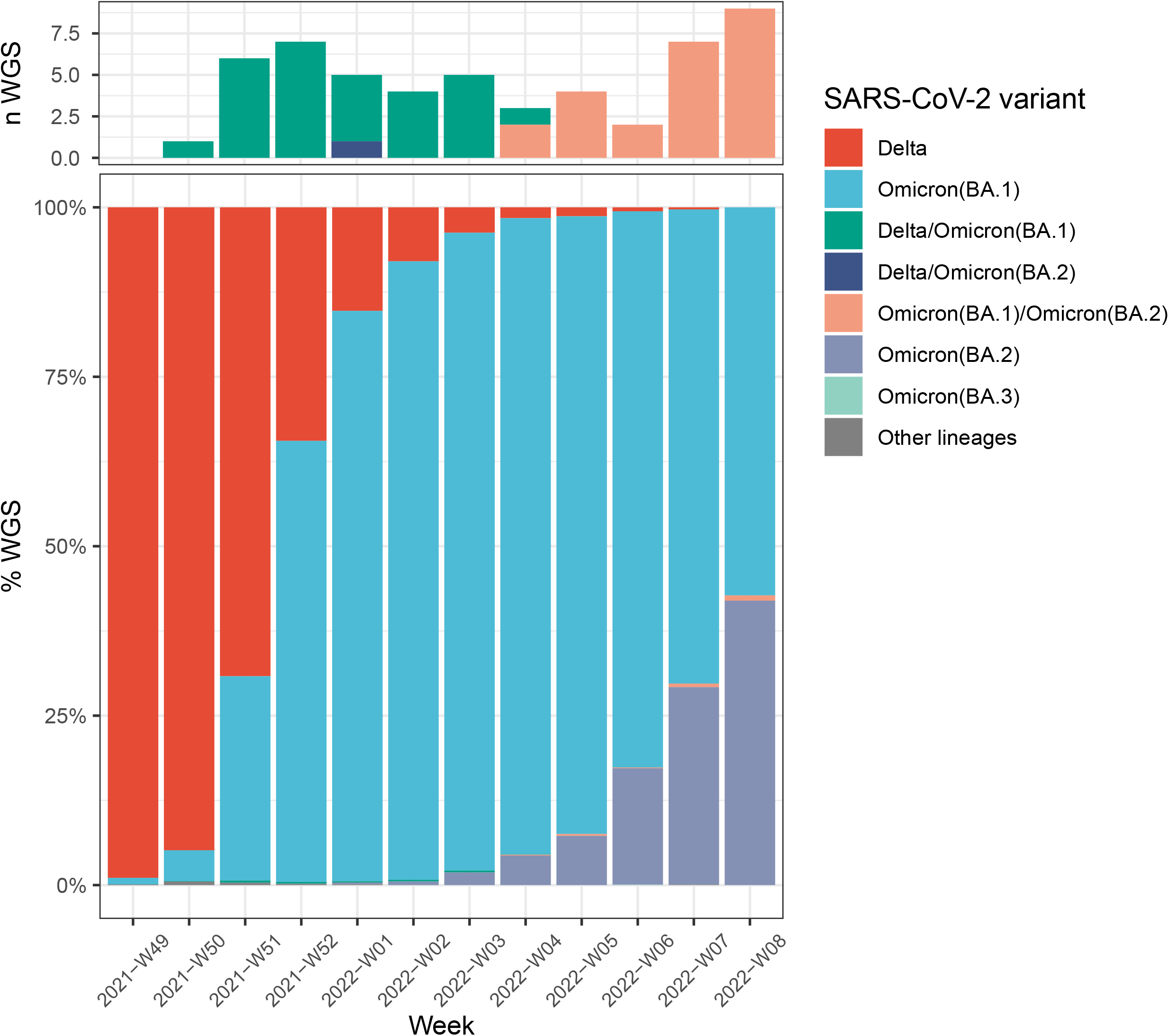
Number of Delta/Omicron (BA.1), Delta/Omicron (BA.2) and BA.1/BA.2 co-infections (A) during Delta, Omicron (BA.1), and Omicron (BA.2) co-circulation in France (B). Colors indicate lineages : Delta (B.1.617.2 and AY.* lineages) in red, Omicron (BA.1) in blue, Omicron (BA.2) in purple, Omicron (BA.3) in light blue, other lineages (including B.1.640.1) in grey. Co-infections between Delta/BA.1 are in green, Delta/BA.2 in dark blue, and BA.1/BA.2 in salmon.

All co-infections were confirmed by visual examination of vcf plots using the lists of specific mutations from our lineage variant database (**Fig S2, Fig S3** and **Fig S4**). Uniform frequencies of lineage-specific mutations along the genome were observed in each sample, except for two samples (“021228537801” among the Delta/BA.1 co-infections and “722000801801” among the BA.1/BA.2 co-infections). In sample 021228537801, frequencies of Delta- and BA.1-specific mutations were reversed between the first and second half of the genome (**Fig S2**). In sample 722000801801, frequencies of BA.1- and BA.2-specific mutations were reversed between the first fourth and rest of the genome (**Fig S4**).

Delta/Omicron (BA.1) co-infections were detected between weeks 50-2021 and 04-2022 (**Fig 4)**. Considering only the period of Delta and Omicron (BA.1) co-circulation at relative frequencies above 1% (Weeks 50-2021 to 05-2022), Delta/Omicron (BA.1) prevalence was estimated at 0.18% (28/15,253; 95 CI : 0.12%-0.26% assuming a binomial distribution). The highest prevalence of Delta/Omicron (BA.1) co-infections was observed in weeks 51 and 52-2021 with 0.31% (6/1,921) and 0.25% (7/2,847), respectively. These 2 weeks were characterized by the highest co-circulation of Delta and Omicron (BA.1) (week 52: 69.2% Delta and 30.1% BA.1; week 52: 34.5% Delta and 65.1% BA.1). BA.1/BA.2 co-infections were detected between weeks 04 and 08-2022 with prevalence reaching 0.78% of the sequences during the highest co-circulation of BA.1 and BA.2 (week 08-2022: 57.3% BA.1 and 42% BA.2). Considering only the period of BA.1 and BA.2 co-circulation at relative frequencies above 1% (Weeks 03 to 08-2022), BA.1/BA.2 prevalence was estimated at 0.26% (24/9,120; 95 CI: 0.17%-0.39%).

### Clinical presentation of Delta/Omicron and BA.1/BA.2 co-infections

To assess the impact of co-infections on clinical presentations, demographic features including age and sex were reported for 13,187 out-patients, 6,242 hospitalized patients, and 803 healthcare workers (**Table 1**). In the three groups, no significant difference was noted between BA.1 and BA.2 infections regarding median age or proportion of men (p>0.05). Therefore, BA.1 and BA.2 cases were grouped into Omicron cases for further analysis. Delta cases were significantly older than Omicron cases for out-patients (p=0.003) and for hospitalized patients (p<0.001). Delta cases were also significantly more predominant in men for hospitalized patients (p<0.001). No difference regarding age or sex was found between Delta and Omicron cases for healthcare workers (p<0.05). Among the three groups, no significant difference in age or sex was found for Delta/Omicron or BA.1/BA.2 co-infections compared to any other type of infection (p>0.05), except for BA.1/BA.2 co-infections significantly reported in younger Delta out-patients (p=0.03) and Delta/Omicron hospitalized patients (p=0.004). ICU admission for hospitalized patients was reported in 4.81%, 15.38%, 1.64%, and 0% of Delta, Delta/Omicron, Omicron and BA.1/BA.2 cases respectively. A binomial regression was tested and after correction by age and sex, ICU admission was significantly associated with Delta (p<0.001) or Delta/Omicron (p<0.001) infection.

**Table 1.** Clinical and demographic data of out-patients, hospitalized patients and healthcare workers infected with Delta, Omicron (BA.1 or BA.2 lineages), or co-infected with Delta/BA.1 or BA.1/BA.2.

| Out-patients |  |  |  |  |  | Hospitalized |  |  |  |  | Healthcare workers |  |  |  |  |
| --- | --- | --- | --- | --- | --- | --- | --- | --- | --- | --- | --- | --- | --- | --- | --- |
|  | Delta<br>(N = 4307) | Delta/Omicron<br>(N = 15) | Omicron<br>(N = 8828) | BA.1/BA.2<br>(N = 21) | Other<br>lineages<br>(N = 16) | Delta<br>(N = 1601) | Delta/<br>Omicron<br>(N = 13) | Omicron<br>(N = 4622) | BA.1/BA.2<br>(N = 3) | Other<br>lineages<br>(N = 3) | Delta<br>(N = 103) | Delta/Omicron<br>(N = 1) | Omicron<br>(N = 699) | BA.1/BA.2<br>(N = 0) | Other<br>lineages<br>(N = 0) |
| **Age** |  |  |  |  |  |  |  |  |  |  |  |  |  |  |  |
| median<br>(IQR) | 39.36<br>(24.01,<br>52.26) | 31.13<br>(19.94,<br>36.09) | 36.93<br>(22.23,<br>51.85) | 22.36<br>(16.66,<br>30.29) | 36.00<br>(27.25,<br>62.54) | 52.57<br>(33.36,<br>73.10) | 66.61<br>(50.79,<br>73.20) | 43.86<br>(26.73,<br>72.86) | 29.03<br>(28.33,<br>30.95) | 53.16<br>(39.23,<br>72.17) | 37.33<br>(28.16,<br>44.24) | 25.10<br>(25.10,<br>25.10) | 32.50<br>(25.74,<br>44.87) | NA | NA |
| **Sex** |  |  |  |  |  |  |  |  |  |  |  |  |  |  |  |
| Male n<br>(%) | 1,990<br>(46.20) | 10<br>(66.67) | 4,035<br>(45.71) | 6<br>(28.57) | 4<br>(25.00) | 852 (53.22) | 7<br>(53.85) | 2,084<br>(45.09) | 2<br>(66.67) | 1<br>(33.33) | 30<br>(29.13) | 0<br>(0.00) | 195<br>(27.90) | NA | NA |
| **Severity** |  |  |  |  |  |  |  |  |  |  |  |  |  |  |  |
| ICU<br>admission<br>(%) | NA | NA | NA | NA | NA | 77 (4.81%) | 2<br>(15.38%) | 76<br>(1.64%) | 0<br>(0.00%) | 0<br>(0.00%) | NA | NA | NA | NA | NA |

## Discussion

The intense co-circulation of distinct SARS-CoV-2 lineages in France since late 2021 gives the unique opportunity to assess the prevalence of Delta/Omicron and BA.1/BA.2 co-infections on a large data set of samples. Herein we found a prevalence of 0.18% and 0.26% for Delta/BA.1 and BA.1/BA.2 co-infections, respectively.

The estimation of the SARS-CoV-2 co-infection prevalence had been performed in 2020 by Liu and colleagues, who reported about 5% of co-infections in the United Arab Emirates. The low genetic diversity of SARS-CoV-2 observed during this period strongly limits the interpretation of this result ^17^. More recently, a study conducted on out-patients in the USA during the same period (November 2021 to February 2022) identified 20 Delta/Omicron co-infections out of 16,386 samples sequenced. The resulting prevalence of about 0.1% for Delta/Omicron is highly consistent with the prevalence found herein. However, no BA.1/BA.2 co-infection was reported by the authors as they specifically looked for Delta/Omicron co-infections using a curated list of mutations ^7^. In contrast, the unbiased co-infection detection script presented herein and validated on mixed Delta:Omicron isolates was defined to detect likely co-infections whatever lineages are present in the sample and can be applied for all SARS-CoV-2 co-infections. Furthermore, rates of detection for specific-Omicron mutations were below 50% in several Delta:Omicron mixes according to the set of primers used for WGS. Thus, the threshold of alternate allele fraction set at 85% by Bolze et al., can miss some co-infection cases and underestimate their prevalence.

With SARS-CoV-2 genetic diversity increasing, amplicon primer sets may lead to drop-out ^18^ and preferential amplification of one lineage over the other in a co-infected sample. Indeed, updates of Midnight and Artic V4 primers into Midnight V2 and Artic V4.1, respectively, were designed to resolve specific Omicron drop-outs. In this study, Artic primers were less biased than Midnight primers for estimation of viral frequencies in experimental mixes, and V4.1 had the best performance in detecting Omicron-specific mutations in mixes with low proportions of Omicron isolates. However, none of the 4 primer sets tested avoided artefactual chimeric sequences in several mixes. Therefore, the detection of co-infections is important to avoid depositing such chimeric sequences in the public repositories. To date (2022-03-18), 2,253 Omicron sequences and 1,156 Delta sequences have been tagged as “Multiple Lineage” in GISAID, including Omicron sequences bearing S:L452R + M:I82T, and may represent Delta/Omicron co-infections, contaminations or genuine recombinants.

The present study brings a first insight into the clinical severity of patients presenting with a co-infection. While Delta/Omicron co-infections were associated with higher rates in ICU admission compared with Omicron infections, no severe forms were noticed in BA.1/BA.2 co-infections. These distinct clinical spectrums need to be further investigated to include the vaccination status of co-infected patients. The clinical characteristics of potential recombinants resulting from a co-infection event ^19^ should also be assessed on a large data set. Among the 53 cases of co-infection identified in the present study, two possible novel recombinants occurring in co-infected samples may have been detected: one potential Delta-BA.1 recombinant in a co-infected sample with unbalanced frequencies of Delta- and Omicron-specific mutations and one potential BA.1-BA.2 recombinant in a co-infected sample with unbalanced frequencies of BA.1- and BA.2-specific mutations. Further investigations, including virus isolation in cell culture, are ongoing to characterize these potential recombinants. In comparison, Bolze et al., identified a Delta-BA.1 recombinant population in 1 out of 20 co-infected cases in the USA ^7^. These data suggest that rates of recombination within co-infected hosts are low (<5%).

This study has several limitations. First, the follow-up duration for the assessment of BA.1/BA.2 co-infection is short, and as BA.2 detection is still increasing in France, the prevalence of BA.1/BA.2 co-infection may be underestimated. In addition, ICU admission and vaccination status information was lacking for some Flash cases, limiting the clinical characterization of co-infected patients. Finally, additional sequencing methods were not tested, such as metagenomics or hybrid capture which are less prone to amplification-bias toward specific lineages ^7^. However, these techniques have lower sensitivity and lower throughput and were therefore not suitable as first-line sequencing methods in our laboratory ^20^.

In conclusion, our findings emphasize the importance of using appropriate experimental and bioinformatic methods for the comprehensive identification of SARS-CoV-2 co-infections. Although these events are rare, SARS-CoV-2 co-infections need to be properly identified as they can lead to the emergence of new variants after a recombination event.

## Acknowledgments

We would like to thank all the members of the GenEPII sequencing platform who contributed to this investigation. We also thank all the laboratories, clinicians and patients involved in this work. This work was carried out within the framework of the French consortium on surveillance and research on infections with emerging pathogens via microbial genomics (consortium relatif à la surveillance et à la recherche sur les infections à pathogènes EMERgents via la GENomique microbienne EMERGEN; https://www.santepubliquefrance.fr/dossiers/coronavirus-covid-19/consortium-emergen)

## Funding

Santé publique France, the French national public health agency.

Caisse nationale d’assurance maladie (Cnam), the national health insurance funds.

“Enhancing Whole Genome Sequencing (WGS) and/or Reverse Transcription Polymerase Chain Reaction (RT-PCR) national infrastructures and capacities to respond to the COVID-19 pandemic in the European Union and European Economic Area” (Grant Agreement ECDC/HERA/2021/007 ECDC. 12221).

## Supplementary material legends

**Table S1: Versions of tools used in the seqmet bio-informatic pipeline**.

**Table S2: Sequencing results for Delta:Omicron mixes**. Analysis results of the vcf based on the covariant list of Delta- and Omicron-specific mutations are compared with results of our unbiased co-infection detection script.

**Figure S1: Representation of variant allele frequency of Delta- and Omicron-specific mutations for Delta:Omicron mixes**. The list of specific mutations is based on co-variant.org. Delta-specific mutations are depicted in red, Omicron-specific mutations in blue and shared mutations are in grey. Horizontal lines at 50% show which mutations are called in the consensus sequence based on the majority rule.

**Figure S2: Representation of variant allele frequency of Delta- and BA1-specific mutations for 28 samples detected as Delta/Omicron (BA.1) co-infections in duplicate sequencing**. The list of specific mutations is based on the seqmet lineage mutation database. Delta-specific mutations are depicted in red, and BA.1-specific mutations in blue. Horizontal lines at 50% show which mutations are called in the consensus sequence based on the majority rule.

**Figure S3: Representation of variant allele frequency of Delta- and BA.2-specific mutations for 1 sample detected as Delta/BA.2 co-infections in duplicate sequencing**. The list of specific mutations is based on the seqmet lineage mutation database. Delta-specific mutations are depicted in red, BA.2-specific mutations in blue grey. Horizontal lines at 50% show which mutations are called in the consensus sequence based on the majority rule.

**Figure S4: Representation of variant allele frequency of BA.1- and BA.2-specific mutations for 24 samples detected as BA.1/BA.2 co-infections in duplicate sequencing**. The list of specific mutations is based on the seqmet lineage mutation database. BA.1-specific mutations are depicted in blue, BA.2-specific mutations in blue grey. Horizontal lines at 50% show which mutations are called in the consensus sequence based on the majority rule.

## Notes

### Competing Interest Statement

The authors have declared no competing interest.

### Funding Statement

Sante publique France, the French national public health agency.
Caisse nationale d assurance maladie (Cnam), the national health insurance funds.
Grant : Enhancing Whole Genome Sequencing (WGS) and/or Reverse Transcription Polymerase Chain Reaction (RT-PCR) national infrastructures and capacities to respond to the COVID-19 pandemic in the European Union and European Economic Area (Grant Agreement ECDC/HERA/2021/007 ECDC 12221)

### Author Declarations

This study was approved by the ethics committee of the Hospices Civils de Lyon (HCL), Lyon, France and registered on the HCL database of RIPHN studies (AGORA Number 41).

## References

1. Tracking SARS-CoV-2 variants. https://www.who.int/health-topics/typhoid/tracking-SARS-CoV-2-variants.

2. Viana, R. et al. Rapid epidemic expansion of the SARS-CoV-2 Omicron variant in southern Africa. Nature (2022) doi:10.1038/s41586-022-04411-y.

3. CoVariants. https://covariants.org/.

4. Enquêtes FlashlJ: évaluation de la circulation des variants du SARS-CoV-2 en France. https://www.santepubliquefrance.fr/etudes-et-enquetes/enquetes-flash-evaluation-de-la-circulation-des-variants-du-sars-cov-2-en-france.

5. Combes, P. et al. Evidence of co-infection during Delta and Omicron variants of concern co-circulation, weeks 49-2021 to 02-2022, France. 2022.03.02.22271694 https://www.medrxiv.org/content/10.1101/2022.03.02.22271694v1 (2022).

6. Rockett, R. J. et al. Co-infection with SARS-COV-2 Omicron and Delta Variants Revealed by Genomic Surveillance. 2022.02.13.22270755 https://www.medrxiv.org/content/10.1101/2022.02.13.22270755v1 (2022).

7. Bolze, A. et al. Evidence for SARS-CoV-2 Delta and Omicron co-infections and recombination. 2022.03.09.22272113 https://www.medrxiv.org/content/10.1101/2022.03.09.22272113v1 (2022).

8. Nyberg, T. et al. Comparative analysis of the risks of hospitalisation and death associated with SARS-CoV-2 omicron (B.1.1.529) and delta (B.1.617.2) variants in England: a cohort study. Lancet Lond. Engl. (2022) doi:10.1016/S0140-6736(22)00462-7.

9. Ulloa, A. C., Buchan, S. A., Daneman, N. & Brown, K. A. Estimates of SARS-CoV-2 Omicron Variant Severity in Ontario, Canada. JAMA (2022) doi:10.1001/jama.2022.2274.

10. Davis, J. J. et al. Analysis of the ARTIC Version 3 and Version 4 SARS-CoV-2 Primers and Their Impact on the Detection of the G142D Amino Acid Substitution in the Spike Protein. Microbiol. Spectr. 9, e0180321 (2021).

11. Liu, T. et al. A benchmarking study of SARS-CoV-2 whole-genome sequencing protocols using COVID-19 patient samples. iScience 24, 102892 (2021).

12. Grubaugh, N. D. et al. An amplicon-based sequencing framework for accurately measuring intrahost virus diversity using PrimalSeq and iVar. Genome Biol. 20, 8 (2019).

13. Kreier, F. Deltacron: the story of the variant that wasn’t. Nature 602, 19 (2022).

14. Fritz, A. et al. Haploflow: strain-resolved de novo assembly of viral genomes. Genome Biol. 22, 212 (2021).

15. https://www.who.int/docs/default-source/coronaviruse/real-time-rt-pcr-assays-for-the-detection-of-sars-cov-2-institut-pasteur-paris.pdf. https://www.who.int/docs/default-source/coronaviruse/real-time-rt-pcr-assays-for-the-detection-of-sars-cov-2-institut-pasteur-paris.pdf.

16. Gaymard, A. et al. Early assessment of diffusion and possible expansion of SARS-CoV-2 Lineage 20I/501Y.V1 (B.1.1.7, variant of concern 202012/01) in France, January to March 2021. Euro Surveill. Bull. Eur. Sur Mal. Transm. Eur. Commun. Dis. Bull. 26, (2021).

17. Genomic epidemiology of SARS-CoV-2 in the UAE reveals novel virus mutation, patterns of co-infection and tissue specific host immune response - PubMed. https://pubmed.ncbi.nlm.nih.gov/34234167/.

18. Sanderson, T. & Barrett, J. C. Variation at Spike position 142 in SARS-CoV-2 Delta genomes is a technical artifact caused by dropout of a sequencing amplicon. 2021.10.14.21264847 https://www.medrxiv.org/content/10.1101/2021.10.14.21264847v2 (2021).

19. Simon-Loriere, E. & Holmes, E. C. Why do RNA viruses recombine? Nat. Rev. Microbiol. 9, 617–626 (2011).

20. Charre, C. et al. Evaluation of NGS-based approaches for SARS-CoV-2 whole genome characterisation. Virus Evol. 6, veaa075 (2020).

