## Supplementary figures and images for "Detection and prevalence of SARS-CoV-2 co-infections during the Omicron variant circulation, France, December 2021 - February 2022"

### Supplementary

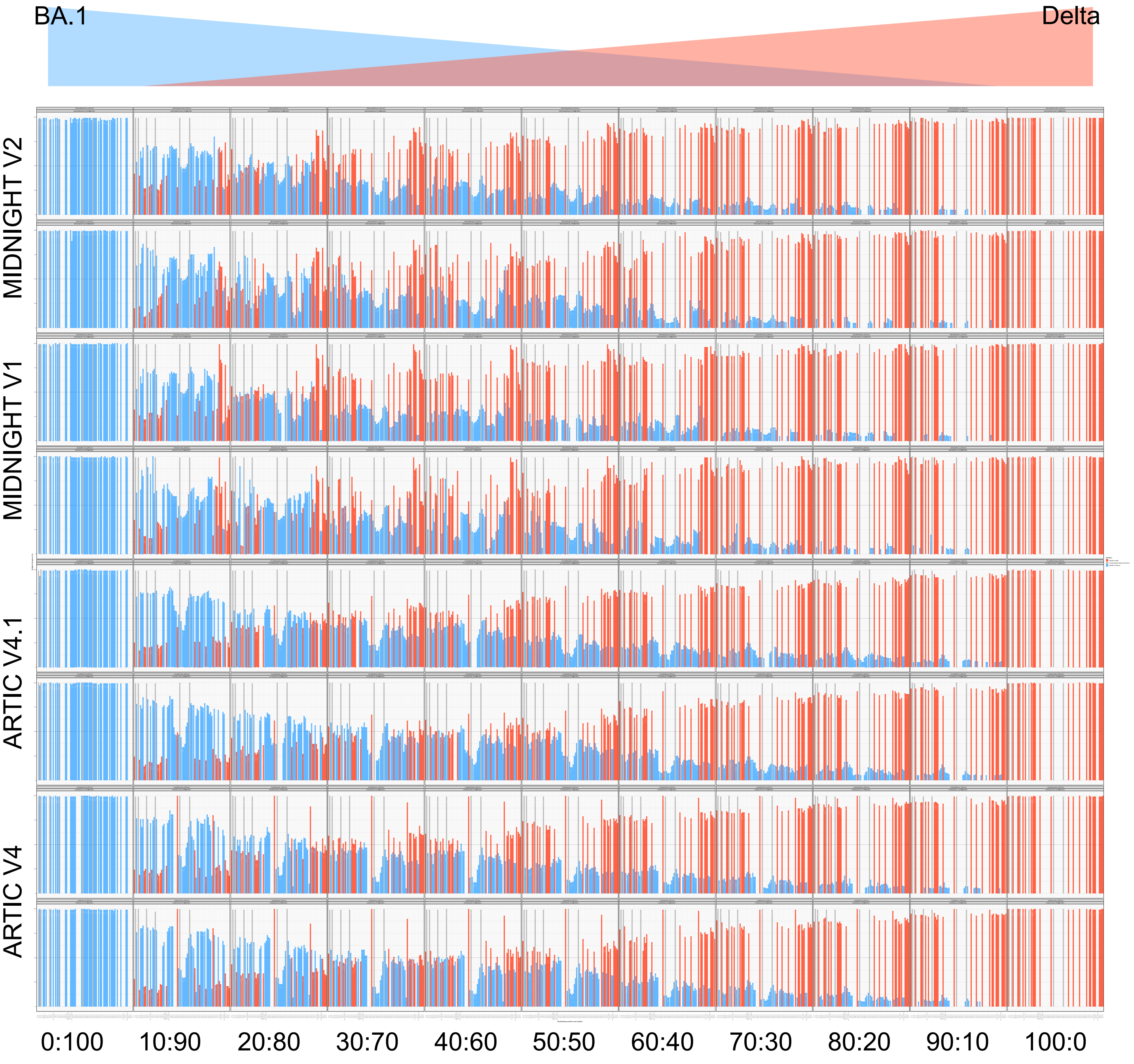

#### Figure S2

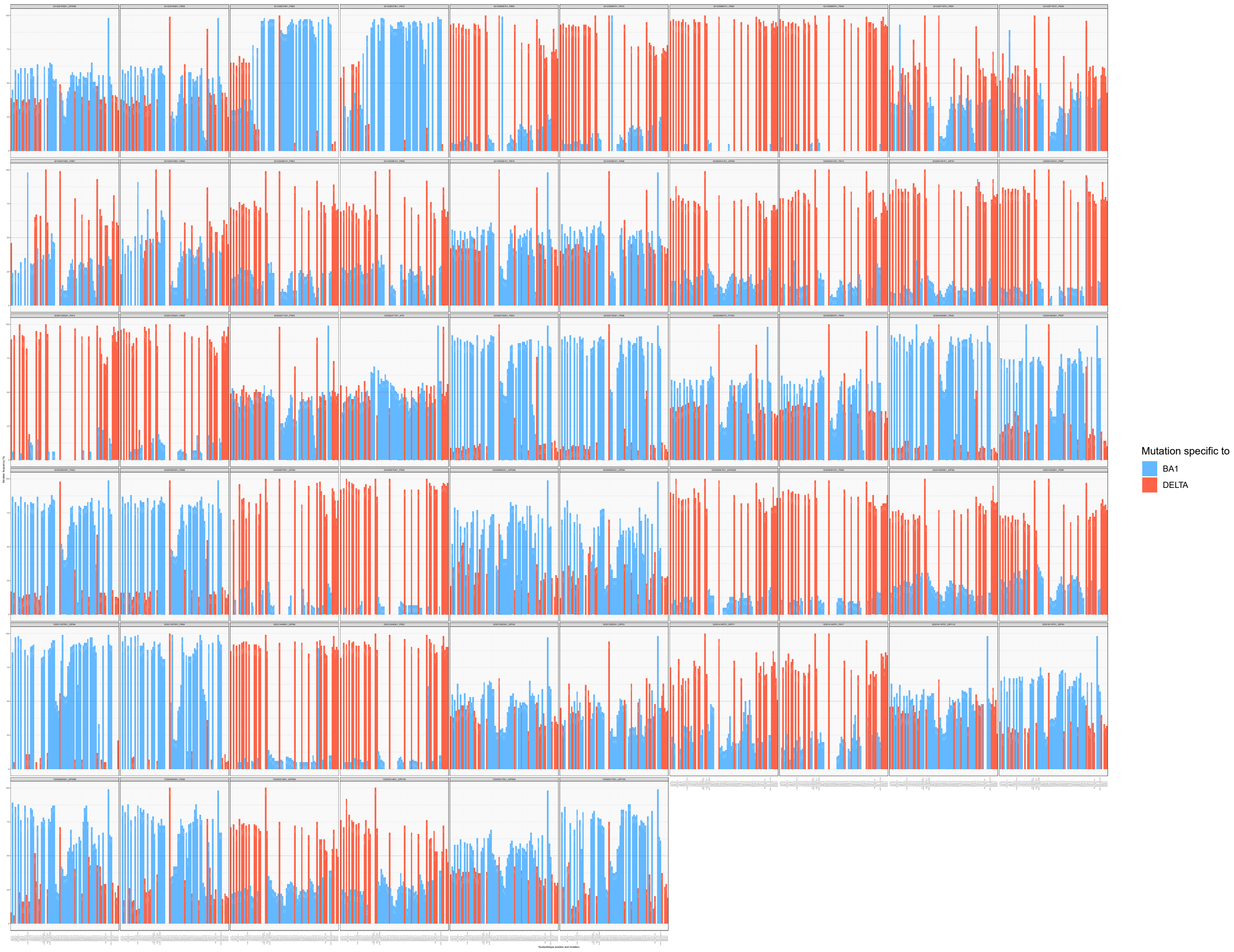

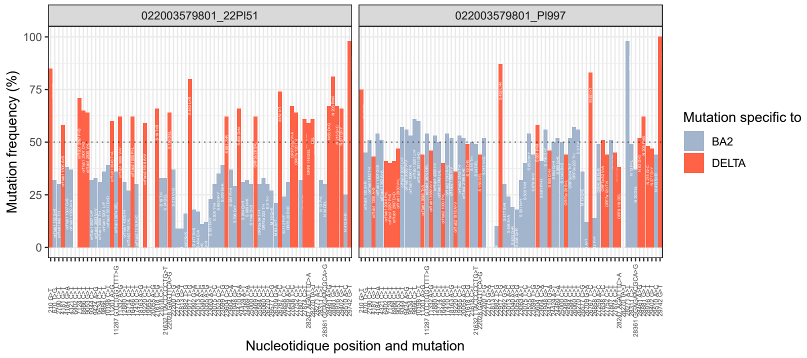

#### Figure S4

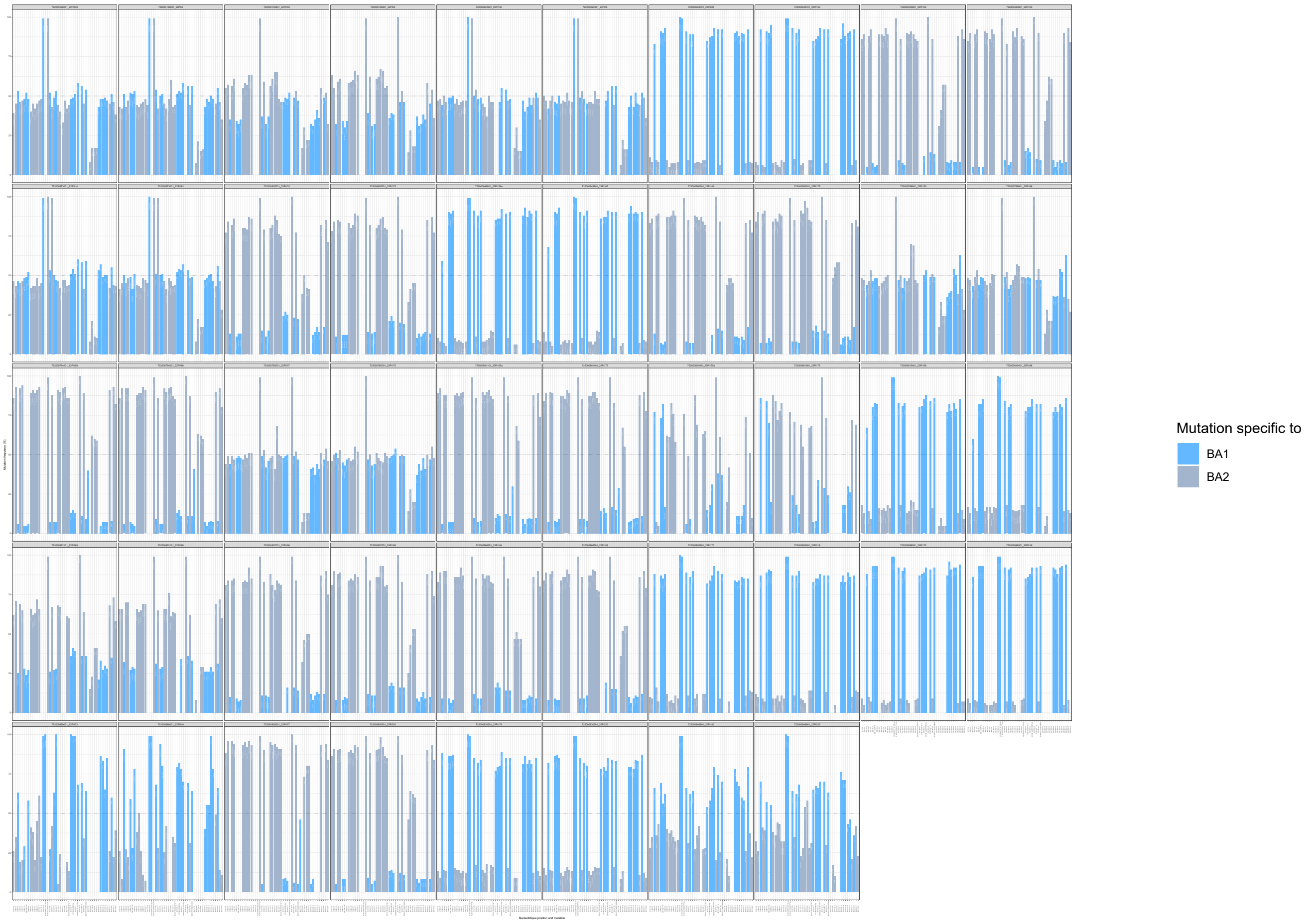
